# Phase II Randomized Study of Short Course Radiotherapy Total Neo-adjuvant Therapy with or without Chlorophyllin in Reducing the Incidence of >/=Grade 2 Acute Toxicity in Advanced Rectal Cancer patients Suitable for Wait and Watch

**DOI:** 10.1101/2024.01.26.24301857

**Authors:** Rahul Krishnatry, Vikram Gota, Debanjan Chakraborty, Vikas Ostwal, Mrs. Sadhana Kannan, Ms Pallavi Rane, Shivakumar Gudi, Mufaddal Kazi, Anant Ramaswamy, Prabhat Bhargava, Purvi Haria, Suman K Ankathi, Munita Bal, Mukta Ramadwar, Ashwin DeSouza, Avnish Saklani, Reena Engineer

**Affiliations:** Department of Radiation Oncology, Tata Memorial Centre, Homi Bhabha National Institute, Mumbai, India; Department of Clinical Pharmacology, ACTREC, Tata Memorial Centre, Homi Bhabha National Institute, Navi Mumbai, India; Department of Epidemiology and Clinical Trials Unit, ACTREC, Tata Memorial Centre, Homi Bhabha National Institute, Navi Mumbai, India; Department of Medical Oncology, Tata Memorial Hospital, Homi Bhabha National Institute, Mumbai, India; Department of Surgical Oncology, Tata Memorial Hospital, Homi Bhabha National Institute, Mumbai, India; Department of Radiology, Tata Memorial Hospital, Homi Bhabha National Institute, Mumbai, India; Department of Pathology, Tata Memorial Hospital, Homi Bhabha National Institute, Mumbai, India

**Author notes:** **Correspondence**- Dr Rahul Krishnatry MD, DNB Professor, Homi Bhabha National Institute (HBNI), Department of Radiation Oncology, Tata Memorial Centre, Dr Ernest Borges Marg, Mumbai, Maharashtra 400012, India., Adress: Room No 1125, 11^th^ floor, HBB Block. **Funding information:** Chlorophyllin and Placebo tablets will be manufactured by IDRS Labs Pvt Ltd. India, and relevant funding will be managed by Bhabha Atomic Research Centre (BARC), along with the run-in costs up to the recruitment of 76 patients for the SCOTCH trial. This funding will cover meetings, central review, treatment planning, toxicity management and operational costs. The study is an investigator-initiated study with the funding agency having no control over the study data, its analysis and presentation. **TRIAL REGISTRATION NUMBER:** CTRI/2023/04/051458 [Registered on: 10/04/2023], ClinicalTrials.gov (NIH): NCT05856305. **WHO trial registration data set version 1.3.1** attached in Appendix 8, S2 file. **PROTOCOL VERSION AND DATE IDENTIFIER:** Protocol Version 2.0 dated 10.02.2023.

**Keywords:** Total Neoadjuvant therapy, Brachytherapy, Chlorophyllin, Wait and watch, Non-operative management, Rectal cancer

## Abstract

**Background:** Total Neoadjuvant treatment (TNT) comprising short-course radiotherapy (SCRT) and induction chemotherapy is one of the standard treatment options for locally advanced rectal cancer (LARC). The addition of localised radiotherapy boost dose using techniques such as brachytherapy can improve local tumour control and organ preservation, in selected good responder patients. Overall increased risk of acute treatment-related toxicity rates with TNT approaches can be a deterrent to compliance, treatment completion and overall outcomes. This phase II study is to evaluate, if the addition of Chlorophyllin to this approach, can reduce the burden of grade 2 or higher acute toxicity – Gastrointestinal (GI)/ Genito-urinary (GU)/ haematological toxicity and the rate of overall complete response (clinical and pathological) in well-selected wait and watch suitable locally advanced rectal cancer patients.

**Aims:** We aim to evaluate the utility of adding chlorophyllin to SCRT-based TNT interdigitated with brachytherapy as applicable in reducing the incidence of grade 2 or higher acute GI/GU/haematological toxicity in advanced rectal cancer along with estimating the rates of complete clinical responses (pathological + clinical) at the end of two years (2-year overall complete response rates). We will be also estimating organ preservation rates, TME-free survival, Disease-free survival, Distant metastasis-free survival, Loco-regional failure-free survival, and Overall survival, along with toxicities and Quality of Life outcomes as secondary objectives.

**Methods:** The study is a 2-arm, phase II, prospective, randomized, double-blind, placebo-controlled superiority study evaluating the clinical outcome - local tumour response, the feasibility of non-operative management (NOM) with hypofractionated dose-escalated radiotherapy, and benefit of Chlorophyllin in reducing toxicity for total neoadjuvant treatment-TNT strategy including short-course radiotherapy and chemotherapy interdigitated with brachytherapy boost for rectal cancer patients. NOM or TME surgery will be followed based on response to NAT as standard treatment in both arms. After accrual and informed consent of eligible LARC patients, there will be: Arm 1 to receive chlorophyllin, and Arm 2 will receive a matching placebo. Permuted block randomisation with a variable block size will be used to randomize 76 (38 in each arm), providing 80% power and a two-sided alpha of 10% to test an absolute reduction in ≥grade 2 GU/GI/Haematological toxicity rates by 30% (from 70% to 40%) with an anticipated dropout of 10%. It will also provide an estimate for NOM and organ preservation success rates. The current sample size is adequate for the estimated overall response rate at 2 years to be 50% compared to pCR of 28% (est. 95% CI: 24% - 32%) as reported in the RAPIDO study. The study started accrual on 04^th^ July 2023 and is currently ongoing.

**Discussion:** We anticipate that with improved logistics of SCRT, better compliance to TNT and improved NOM rates with endorectal brachytherapy boost could be achieved with Chylorophyllin by ameliorating acute treatment-related GI/GU /Haematological toxicity rates. Improved NOM rates and lesser toxicity would result in superior QoL and improved therapeutic ratio compared to the usual high toxicity noticed in standard SCRT-based TNT strategies and TME employed globally.

## 1. INTRODUCTION

Historically, neoadjuvant chemoradiation (CTRT) followed by total mesorectal excision +/- adjuvant chemotherapy based on histopathological parameters has been standard of care in locally advanced rectal cancer, with a 15-27% pathological complete response [1][2]. However, significant rates of postoperative complications (18% to 38%), mortality (around 3%), and especially poor QoL related to permanent colostomy and bowel morbidity related to APR in lower rectal tumours remain a challenge [3].[4]. Because of this, Non-operative management (NOM) strategies after neoadjuvant chemoradiation therapy have been on the rise [5]. Complete clinical response (cCR), like pCR, turning into fruitful Non-operative management (NOM) is achieved in 15-25% with the LCRT dose of 45-50Gy [6]. Prospective dose-response studies showed a direct relationship between the escalated radiotherapy dose and local tumour showing that, >100 BED Gy10 is needed for complete response in >80% of tumours [7-9]. Currently, NOM management is one of the standard treatment options in suitable patients as per various international guidelines like NCCN [10].

Despite improved local control with radiotherapy and surgery, many patients (20-25%) develop distant metastasis even after adjuvant chemotherapy, mostly due to delayed timing of chemotherapy and poor compliance, low treatment completion rates of up to 55% [11][12]. Recently, total neoadjuvant treatment (TNT) schedules have been developed where radiotherapy and chemotherapy are completely delivered before surgery or a decision for NOM. One such approach is (RAPIDO study) SCRT followed by induction chemotherapy [13], demonstrating, lower rates of disease-related treatment failure and reduced cumulative probability of distant metastasis compared to standard CTRT. Apart from sparing radiotherapy recourses (1/5^th^), the pathological CR was double (28%) but no significant NOM outcomes were available as that was not looked for. Although the overall toxicity rates are balanced irrespective of the sequencing of treatments, the rates are still higher and need to be addressed. The commonest grade 3 or higher adverse event during TNT was diarrhoea (18%) with any grade 3 or higher toxicity at 38%. Overall, the per-patient highest toxicity of >grade 2 GI/GU/Haematological CTCAE v 5 is expected to affect about 70% of patients warranting dose reduction in 44% and premature cessation at about 15%.

A recent non-clinical study has shown the utility of Sodium-copper-chlorophyllin (CHL) as a protective agent against cytotoxicity, being a good free radical scavenger- in mice receiving whole-body irradiation of 7.5 Gray radiation dose. Chlorophyllin conferred protection against GI toxicity such as loss of absorptive surface, reduction in villi length and irregular epithelial alignment, and prevented radiation-induced pulmonary atypia [14]. Also, in phase 1, single ascending dose study, a 3 g/day dose of chlorophyllin was found to be safe [unpublished data]. After pre-clinical model validations, Chlorophyllin was the obvious choice when exploring non-invasive, easily available, and safe options to address this treatment-related toxicity. It is a phytopharmaceutical semi-synthetic water-soluble mixture of sodium copper salts derived from green plant pigment, chlorophyll. It is also used for many uses like food colorant, prevention of body odour in geriatric patients, enhanced wound healing, antibacterial action, prevention of cancer in the high-risk population exposed to hepatocarcinogen aflatoxin B1, treatment of faecal incontinence etc. [15]. Studies have shown that CHL has immunostimulatory, anti-inflammatory and antiviral effects too [14][16]. It increases the expression of a transcription factor (protein) Nrf2 which improves lymphocyte survival and enables efficient detoxification after exposure to radiation, along with delayed microtubule polymerization and mitotic spindling - also mitigating radiation damage. It also ameliorates radiation-induced toxicity in normal hematopoietic tissues and epithelial cells [16]. However, chlorophyllin accentuates the radiation-induced killing of human breast cancer cells in xenograft tumour-bearing SCID mice [17]. It was found to be well tolerated up to an acute oral dose of 5000 mg/kg body weight in mice and rats. A 1000 mg/kg body weight oral dose repeated for 28 consecutive days was also well tolerated in rodents.

With this study, we hypothesize that the addition of chlorophyllin will decrease acute grade 2 or higher GI/GU/Haematological toxicity (CTCAE v 5) rates by 30% (from the current 70% to 40%) among the patients of locally advanced rectal cancer, receiving chlorophyllin versus those who will not, both undergoing SCRT based TNT schedule interdigitated with endorectal brachytherapy-based TNT treatment as a part of the NOM strategy in well-selected patients as per the international standard criteria (IWWD) [18][19]. We would also estimate the NOM success rates and overall complete response rates (cCR and pCR) at 2 years from diagnosis in the overall cohort irrespective of Chlorophyllin status, anticipating the CR overall will reach 50% at 2 years compared to 28% pCR depicted in RAPIDO study.

## 2. METHODOLOGY

### Primary Objective

To evaluate the utility of adding chlorophyllin to SCRT and endorectal brachytherapy-based TNT in reducing the incidence of grade 2 or higher acute GI/GU/haematological toxicity in advanced rectal cancer, while assessing the overall local response rates.

### Secondary Objectives

- To estimate the 2-year overall complete response rates (clinical or pathological) in the whole cohort and if there is any difference between chlorophyllin and control arms.
- To estimate the 2-year organ preservation rates, TME-free survival and if any difference between chlorophyllin and control arms.
- To estimate the 2-year disease-free survival, Distant metastasis-free survival, loco-regional failure-free survival, and overall survival rates in the whole cohort, and if any difference between two arms and between patients with successful NOM versus others.
- To compare treatment-related early and late toxicities (≥grade 2 CTCAE v5) for two years between the groups as (3).
- To estimate surgical complications based on Clavien-Dindo classification.
- To estimate and compare HRQOL (EORTC QOL-C30, CR 29, PRT, SH 22), and LARS scores and between various groups as (3).
- To estimate direct cost benefit with reduction in toxicity.
- To study the tumour volume reduction kinetics and radiotherapy doses and probability of successful NOM outcomes.

#### DESIGN AND SETTING

The study is a 2-arm, phase II, prospective, randomised, double-blind, placebo-controlled superiority study evaluating the probability of successful NOM in a setting of SCRT-based TNT and the benefit of Chlorophyllin in reducing acute treatment-related toxicity in locally advanced rectal cancer.

#### SAMPLE SIZE CALCULATIONS

A sample size of 68 subjects (34 in each arm) with 1:1 randomisation (Chlorophyllin versus Placebo) provides 80% power with a two-sided alpha level of 10% to test an absolute reduction in the primary endpoint of grade 2 or worse GU/GI/Haematological toxicity rates by 30%. The toxicity rate in the Placebo arm is assumed to be 70% (13) and chlorophyllin is expected to decrease it to 40%. To compensate for an anticipated dropout of 10% a total of 76 (38 in each arm) will be randomised in the study. This will also provide an adequate confidence level to estimate the NOM success rates measured by complete clinical response (cCR) at 2 years from completion of TNT in the overall cohort irrespective of Chlorophyllin status. It is expected that cCR at 2 years would be 50%, as compared to pCR of 28% (est. 95% CI: 24% - 32%) as reported in the RAPIDO study. The sample size is expected to produce a two-sided 95% confidence interval with a width equal to 0.25 when the cCR is expected to be around 50% with a lower limit of 37.5% which will exclude the estimated upper limit of around 32% in the RAPIDO study.

#### CONSENTING, RECRUITMENT, RANDOMIZATION, ALLOCATION AND BLINDING

All incoming out-patients with LARC as per the eligibility criteria in **Table 1**, will be screened until the target population is achieved (76 patients). After the agreement of the patient to participate and informed consent (by reading the validated ICF in a language understood by the patient) following good clinical practice and ethical codes, randomization will be done by the study statisticians, centrally **-**using a computer-generated random sequence. Permuted block randomization with variable block size, maintained at CRS, Tata Memorial Centre will be employed. Being a double-blinded placebo-controlled trial, patients in one arm will receive Sodium Copper Chlorophyllin 750 mg tablets for oral administration whereas patients in the control arm will receive an identical matching placebo with unique ID No. given by the manufacturer (CONSORT; Figure 1). These numbers will be available with an unblended statistician along with information about whether the ‘numbered bottle’ contains a ‘test drug’ or ‘placebo.’ The study pharmacist will dispense bottles as instructed by an unblinded statistician. The treating physicians, patients and the blinded statistician will remain blinded to the treatment.

**Table 1.** Inclusion & Exclusion Criteria for the study.

| Inclusion Criteria | Exclusion Criteria |
| --- | --- |
| 1. Age ≥ 18 years.<br>2. Histologically confirmed diagnosis of adenocarcinoma of the rectum.<br>3. Clinical Stage II/III (T2-3, 4b: adherent to prostate, SV or post vagina but not grossly invading, N0-2) based on MRI.<br>4. Non-circumferential tumours with craniocaudal length ≤ 7 cm<br>5. The tumours of the lower rectum, or starting up to 7 cm from the anal verge.<br>6. No evidence of distant metastases on CT Chest and Abdomen.<br>7. No prior pelvic radiation therapy<br>8. No prior chemotherapy or surgery for rectal cancer<br>9. ECOG Performance status 0-2<br>10. Patients must read, agree to, and sign a statement of Informed Consent prior to participation in this study.<br>11. Eligible to receive one of the options of standard neoadjuvant chemotherapy as determined by the medical oncologist team.<br>a. ANC > 1.5 cells/mm<sup>3</sup>, HGB > 8.0 gm/dl, PLT > 150,000/mm<sup>3</sup>,<br>b. Total bilirubin ≤ 1.5 x ULN (except in patients with Gilbert's Syndrome who must have total bilirubin ≤ 3.0 x ULN), AST ≤ 3 x ULN, ALT ≤ 3 x ULN. | 1. Signet or mucinous histology cancer of rectum<br>2. Recurrent rectal cancer, or previous pelvic radiotherapy<br>3. Primary unresectable rectal cancer.<br>4. Creatinine level greater than 1.5 times the upper limit of normal.<br>5. Patients who are unable to undergo an MRI.<br>6. Patients with a history of any arterial thrombotic event within the past 6 months. This includes angina (stable or unstable), MI, TIA, or CVA.<br>7. Ulcerative colitis or any other histologically confirmed inflammatory bowel disease.<br>8. Patients with a history of venous thrombotic episodes such as deep venous thrombosis, and pulmonary embolism occurring more than 6 months prior to enrolment may be considered for protocol participation, provided they are on stable doses of anticoagulant therapy. Similarly, patients who are anticoagulated for atrial fibrillation or other conditions may participate, provided they are on stable doses of anticoagulant therapy.<br>9. Patients with any other concurrent medical or psychiatric condition or disease which, in the investigator's judgement, would make them inappropriate candidates for entry into this study.<br>10. Poor reliability for follow-up.<br>11. Ineligible as per inclusion and eligibility criteria. |

**Figure 1.**
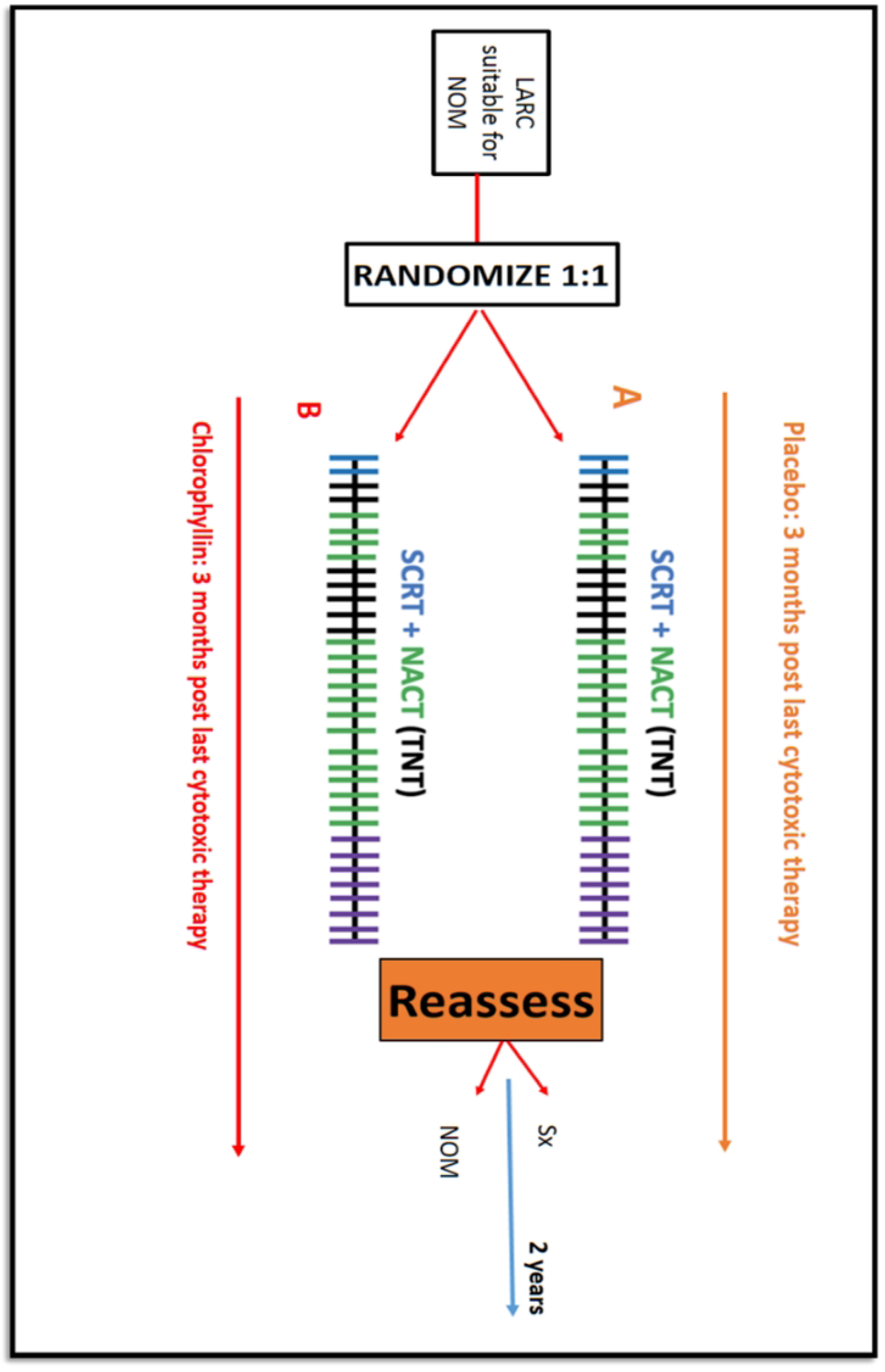
Schedule of enrolment, interventions, and assessments. CONSORT Diagram.

#### INTERVENTIONS

Standard treatment protocol comprising short-course radiation followed by neoadjuvant chemotherapy followed by NOM or Surgery will be applicable in both study arms. The addition of chlorophyll and placebo is the only intervention planned, for this placebo-controlled study.

#### STUDY DRUG – CHLOROPHYLLIN (CHL) AND PLACEBO

The CHL would be stored in a cool, dry place. Each pack of CHL 750 mg/placebo will contain 30 tablets of CHL/placebo tablets-prepared by the Clinical Research Secretariat team at Tata Memorial Centre. It will be administered 750 mg orally once daily in the morning before food, starting 2 weeks before EBRT and continuing for up to 3 months after the last dose of cytotoxic therapy as chemotherapy or radiotherapy. Instructions including dose timing, storage, consumption, and what to do in the event of a missed dose will be explained to the patient by the study nursing staff/ doctor, along with maintaining a pill diary for the patient. Placebo is a chemically inert, stable, and biologically safe compound containing colourant, like the appearance of CHL tablets. It may or may not cause green stools. However, it satisfies all the requirements of a ‘matching placebo’.

##### EXTERNAL BEAM RADIOTHERAPY (EBRT)

All patients will undergo CT-based radiotherapy planning using rotational arc-based intensity-modulated radiotherapy (IMRT) with Target volume contouring will be based on Valentini et al consensus guidelines [20]. Segmentation of Organs at Risk (OAR) will be done according to UK-BFCO Consensus guidelines. The GTV: primary or gross nodes may be boosted to doses of 30 Gray in 5# while the dose to CTV pelvis will be up to 25 Gy in 5 fractions. The plan evaluation for the coverage and OAR constraints will be done as per international anorectal radiotherapy standard guidelines and will be accepted with up to 10% individual patient variation [21]. The deviation from predetermined dose constraints will be reported as acceptable (10%), minor (10-20%) and major deviations (>20%).

##### CHEMOTHERAPY

All patients will be assessed for fitness for chemotherapy by Gastrointestinal - Medical Oncologist and will receive 9 cycles of FOLFOX, administered every 2 Weekly (total 18 weeks) or CAPE OX, every 3 weeks for 6 cycles (18 weeks) or Single 5 FU every 2-weeks for 18 weeks, as per the latest international guidelines such as NCCN v 2022 [10], with recommended laboratory investigations, and clinical review 1 week before and 2 weeks after each chemotherapy cycle. Intervention/dose reduction or cessation of chemotherapy will be planned according to patient tolerances and standard guidelines. Chemotherapy will start 1-4 weeks after the SCRT and will be interdigitated with a brachytherapy schedule - after 1-2 cycles of chemotherapy / 6-12 weeks from SCRT completion, or whenever the clinical response to the primary tumour is found suitable for boost during a total 18 weeks of chemotherapy. An interim gap of a minimum 1-week will be maintained.

#### BRACHYTHERAPY BOOST

At 6-12 weeks after SCRT, periodic assessment by Digital Rectal Examination (DRE) for denoting tumour volume reduction and suitability of brachytherapy boost will be done, during ongoing induction chemotherapy after EBRT. Weekly MRI-guided brachytherapy planning (T2, Axial) followed by intraluminal brachytherapy will be performed as per departmental standard protocol using an indigenously developed endoluminal applicator, to doses of 7-10 Gy/ fraction aiming at a target cumulative BED of 95 Gy (a/b 10) as the maximum dose.

#### ASSESSMENTS POST TNT

The first response assessment comprising of DRE, sigmoidoscopy, and pelvis MRI, will be done at 8-12 weeks (ideally at 6 weeks) after TNT completion for further decision on NOM versus surgery based on clinic-radiological response. A complete clinical response or near complete response without regrowth usually are selected for the NOM-Watch and Wait strategy. [19] The definition of cCR/ ncCR will be decided as per standard Watch and Wait Database (IWWD) group consensus definitions. If disease progression occurs during TNT, further management will be done as per standard recommendations.

Toxicity assessment will be done starting from the time just before initiating SCRT (baseline), then weekly during radiation treatment, and on each scheduled follow-up. Acute toxicity will be considered till 3 months from the last treatment (chemotherapy for NOM and surgery for operated cases) using CTCAE V.5.0 criteria. After 3 months from the last cytotoxic treatment (NOM eligible) or surgery, any toxicity will be deemed as late toxicity and will be documented as aforesaid criteria. Any unplanned interim visit/ hospital admission will be also recorded including toxicity grade at that time using the same assessment tools. QoL including functional outcome will be studied by serving patients with a Questionnaire of EORTC QLQ C30 (Cancer), CR (Colo-rectal) 29, PRT-20 (proctitis), SH (Sexual) 22- (Hindi, Marathi and Bengali versions, validated translations), LARS score and IPSS at start of the treatment, first assessment at 8 to 16 weeks post-TNT completion and thereafter 6 monthly for 2-3 years (+/-1 month) Patient-reported outcomes will be analysed after obtaining the responses. Intra-institutional ePRO (Patient-reported outcomes) forms will be tested as per feasibility, along with Google forms for better adherence of patients.

##### ANALYSIS

Descriptive analyses will be performed to describe the study population. Chi-square tests and unpaired t-tests will be computed to compare relevant sociodemographic and illness-related variables. Simple ratios and numbers will be used to express and compare percentages as relevant. The primary endpoint toxicity and other ratios will be compared using Fisher’s exact test. All primary analyses will be based on intention-to-treat. A per-protocol analysis will be performed as a secondary analysis. Curves for disease-related treatment failure and overall survival-related outcomes will be constructed using the method of Kaplan and Meier. Cumulative incidence of local recurrence or other recurrences will be computed accounting for death as a competing risk. Differences in endpoints will be tested with the log-rank test. Hazard ratios and 95% confidence intervals (CI) will be computed using Cox regression. All tests will be two-sided. The statistical analysis will be carried out by a ‘blinded’ statistician who is blind to study arms. Univariate and multivariate analyses will be performed to examine relationships between QoL scores and the sociodemographic and illness-related variables.

## DISCUSSION

As neoadjuvant chemoradiation is the cornerstone modality in managing LARC, standard long-course neoadjuvant chemoradiation (25 fractions over 5 weeks) approximately 15-27% will achieve a pathologic complete response (pCR) with excellent long-term outcomes, as noted in the literature. But there is already a handful of evidence reflecting that the combined modality therapy in the approach of TNT-total neo-adjuvant treatment including short-course RT and Induction chemotherapy for locally advanced rectal carcinoma is associated with an increase in DFS, along with increased pCR and compliance rate. However, the toxicities can impede compliance and patient quality of life. Also, provisioning double the pCR, presents a significant opportunity for successful NOM with further strengthening using endorectal brachytherapy-based local radiotherapy dose escalation.

High perioperative morbidity, with a poor QoL and bowel function, often with a permanent stoma in lower rectal tumours makes NOM a desired strategy in lower-mid rectum patients. Improved tolerance of TNT treatment with decreased acute toxicity during treatment and the feasibility of higher success rates of NOM would be a win-win for so many deserving patients. Being locally advanced cancers, the TNT approach would also help decrease the risk of long-term failures due to distant metastasis which may be a risk in NOM management in these patients otherwise. The radioprotective, anti-cytotoxic and regenerative properties of chlorophyllin, have already been noted in pre-clinical studies and phase 1 ascending dose studies with minimal side effects and hence we chose to see in our study if chlorophyllin can reduce the acute gastrointestinal/ genitourinary and haematological toxicity related to treatment, in a setting of short course EBRT based TNT approach (+/- endorectal brachytherapy boost in selected group). The 2-year overall local tumour response and organ preservation rates, with this approach of SCRT-based TNT with endorectal boost, will be assessed too as a key objective with adequate sample size.

## 4. CONCLUSION

In the SCOTCH study, we want to explore, if adding chlorophyllin to the neoadjuvant treatment of locally advanced rectal cancer can ameliorate the high rate of acute treatment-related GI/ GU and haematological toxicities, to aim realistically at an increased opportunity of organ preservation through hypofractionated radiotherapy and long duration of systemic chemotherapy via TNT, with a high QoL and better therapeutic ratio.

## Data Availability

The study is ongoing and data-sharing requests can be entertained based on the guidance of approved protocol limits and Ethics committee approval, as applicable.

N.A.

## ACCESS TO DATA/ SHARING POLICY

Anonymised individual patient data can be requested formally via data monitoring unit and afterwards, with proper permission, accessed through principal investigator, Dr Rahul Krishnatry – at for research purpose only. The data will not be shared with study sponsors who do not have any role in the study design, collection, management, analysis, interpretation, and data writing. The final authority for the data sharing and previously mentioned objectives lies with the Principal Investigator and Co-Principal Investigator themselves.

## ROLE OF STUDY SPONSOR AND FUNDERS, IF ANY, IN STUDY DESIGN

This is an investigator-initiated, single-centre study. The design, conduct and interpretation of academic publications will remain the sole right of the investigators. The financial funding source had no role in the design of this study and will not have any role during its execution, analyses, interpretation of the data, or decision to submit the results. Members have specific designated roles for the design and conduct of the study.

**Principal Investigator and Research Physician**: Dr Rahul Krishnatry

**Co-Principal Investigator:** Dr Vikram Gota

**Trial Management Committee (TMC)-** Dr Rahul Krishnatry, Dr Vikram Gota

